# Gain-of-function MARK4 variant associates with pediatric neurodevelopmental disorder and dysmorphism

**DOI:** 10.1101/2023.08.25.23294399

**Authors:** Simran Samra, Mehul Sharma, Maryam Vaseghi-Shanjani, Kate L. Del Bel, Loryn Byres, Susan Lin, Joshua Dalmann, Areesha Salman, Jill Mwenifumbo, Bhavi P. Modi, Catherine M. Biggs, Cyrus Boelman, Lorne A. Clarke, Anna Lehman, the CAUSES Study, Stuart E. Turvey

## Abstract

Microtubule affinity regulating kinase 4 (MARK4) is a serine/threonine kinase that plays a key role in tau phosphorylation and regulation of the mTOR pathway. Abnormal tau phosphorylation and dysregulation of the mTOR pathway are implicated in neurodegenerative and neurodevelopmental disorders.

Here, we report a novel gain-of-function variant in *MARK4* in two siblings with childhood-onset neurodevelopmental disability and dysmorphic features.

The siblings carry a germline heterozygous missense *MARK4* variant c.604T>C; p.Phe202Leu, located in the catalytic domain of the kinase, which they inherited from their unaffected, somatic mosaic mother. Functional studies show that this amino acid substitution has no impact on protein expression but instead increases the ability of MARK4 to phosphorylate tau isoforms found in the fetal and adult brain. The *MARK4* variant also increases phosphorylation of ribosomal protein S6, indicating upregulation of the mTORC1 pathway.

This is the first study to link a germline monoallelic *MARK4* variant to a childhood-onset neurodevelopmental disorder characterized by global developmental delay, intellectual disability, behavioral abnormalities, and dysmorphic features.

## Introduction

Tau is a microtubule-associated protein, well-known for its role in promoting microtubule assembly and stabilization throughout all stages of development. Post-translational modifications of tau, such as hyperphosphorylation, disrupt microtubule binding and promote tau misfolding, leading to the formation of intracellular accumulations of abnormal tau filaments seen in neurodegenerative disorders, collectively known as tauopathies.^1-3^ Tau also plays a central role in the developing brain^4^; hyperphosphorylated tau and aggregates of hyperphosphorylated tau have been observed in neurodevelopmental disorders, especially those with upregulated activity of the mammalian target of rapamycin (mTOR) signaling pathway.^5^

*MARK4* (MIM: 606495) encodes microtubule affinity regulating kinase 4, a member of the MARK protein family, which consists of four serine/threonine kinases (MARK1–4).^6^ MARK4 is predominantly expressed in the brain and its downstream targets include tau and mTOR Complex 1 (mTORC1).^6,7^ Thus, the kinase helps regulate microtubule stability, cell growth, and intracellular signaling. Overexpression of MARK4 in rat hippocampal neurons leads to tau hyperphosphorylation.^8^ A *de novo* variant resulting in a double amino acid change in the common docking domain of MARK4 p.Gly316_Glu317delinsAsp (designated ΔG316E317D), causes increased tau phosphorylation at a specific serine residue (S262) and is associated with early-onset Alzheimer’s disease.^9,10^

To date, *MARK4* variants have not been directly associated with any childhood-onset neurological phenotype. Here, we describe a novel neurodevelopmental disorder associated with a germline heterozygous *MARK4* variant (c.604T>C; p.Phe202Leu) found in two siblings. Both siblings show a highly consistent phenotype of developmental delay, intellectual disability, behavioral abnormalities, and dysmorphic features. The variant is in the catalytic domain of MARK4 and results in gain-of-function leading to hyperphosphorylation of tau isoforms in the fetal and adult brain, and upregulation of the mTORC1 pathway.

## Materials and methods

### Patients

Written informed consent was obtained according to the Declaration of Helsinki for the two brothers (Patients 1 and 2) and their parents for specimen collection, genetic analysis, and publication of photographs and results. Research study protocols were approved by The University of British Columbia Clinical Research Ethics Board (H15-00092 and H18-02853).

### Exome sequencing, analysis, and variant validation

Genomic DNA was extracted from blood samples using standard procedures. Trio-based whole exome sequencing was undertaken with DNA samples from Patient 1 and parents (Supplementary material). Sanger sequencing was performed with DNA samples from both patients and parents (Supplementary material). Primer sequences are detailed in Supplementary Table 1.

### Generation and expression of MARK4 and tau proteins

Plasmids expressing MARK4 and tau proteins were generated through site-directed mutagenesis and cloning for transfection purposes (Supplementary material). A turbo green fluorescent protein (tGFP) tag was included in all MARK4 plasmids while a OFPSpark (a red fluorescent protein) tag was included in all tau plasmids. Gene expression was induced transiently in HEK293 cells using lipofectamine (Supplementary material).

### Immunoblotting

Expression of MARK4^wt^ and MARK4 variant proteins was quantified by immunoblotting. Transfected cells were harvested after 24 hours and lysed (Supplementary material). Cell lysates were separated using gel electrophoresis and transferred onto membranes. Membranes were blocked, incubated with antibodies, and scanned.

### Flow cytometry

Phosphorylation of S6 and tau proteins was quantified in transfected HEK293 cells using phospho-flow cytometry. Twenty-four hours after transfection, cells were harvested, fixed, and permeabilized (Supplementary material). Cells were stained with intracellular flow antibodies and acquired on a BD LSRFortessa™ Cell Analyzer and analyzed using FlowJo.

### Statistical analysis

Statistical analysis was performed using GraphPad Prism 9 Software. Groups were compared using one-way ANOVA with Dunnet’s multiple comparison test or two-way ANOVA with Tukey’s multiple comparison test. \**P* < 0.05; \*\**P* < 0.01; \*\*\**P* < 0.001; \*\*\*\**P* < 0.0001.

## Results

### Novel *MARK4* variant in siblings with childhood-onset neurological symptoms and dysmorphic features

Patient 1 (designated II-1 on the family pedigree) is a male (ages 17-22 years) born to healthy, non-consanguineous parents. He has a longstanding history of developmental delay with significant intellectual disability, early failure to thrive with hypoglycemia, and seizure disorder of presumed genetic but otherwise unexplained etiology (Supplementary Table 2). As a child, he was also diagnosed with autism and attention deficit hyperactivity disorder. Patient 2 (designated II-2 on the family pedigree) is also a male (ages 11-16 years) and the younger brother of Patient 1. Patient 2 has a similar phenotype to his brother with developmental delay, moderate intellectual disability, and behavioral challenges (Case reports and Table 2 in Supplementary material). Both patients are short in stature, with Patient 1 having a body mass index consistently below the 3^rd^ percentile for age. Physical examination of the siblings revealed that they share distinct dysmorphic features; including hypertelorism, ptosis, a shortened columella with coarseness to the alae nasi, pectus excavatum, and prominent fetal pads. Patient 2 has abnormal hearing loss while Patient 1 has no hearing anomalies.

Both patients were suspected of having an underlying single-gene disease however could not be diagnosed with an existing genetic syndrome. Trio-based whole exome sequencing (Patient 1 and both parents) was undertaken to identify a potential genetic etiology. Patient 1 was found to carry a novel heterozygous missense *MARK4* variant (NM_001199867.1:c.604T>C; NP_001186796.1:p.Phe202Leu (F202L)) (Supplementary Table 3). *In silico* bioinformatic tools provided mixed predictions of impact (e.g., high combined annotation dependent depletion (v1.6) score of 27 and low rare exome variant ensemble learner score of 0.313).^11,12^ The mother is mosaic for the MARK4 p.F202L variant but is healthy. Eleven of the 97 sequence reads from the maternal exome sequencing data had the variant while 86 reads were MARK4^wt^ (i.e., ∼10% mosaicism). Sanger sequencing confirmed the variant in Patient 1 and identified the same variant in Patient 2, confirming the variant segregated with disease within the family (Fig. 1A and B).

**Figure 1.**
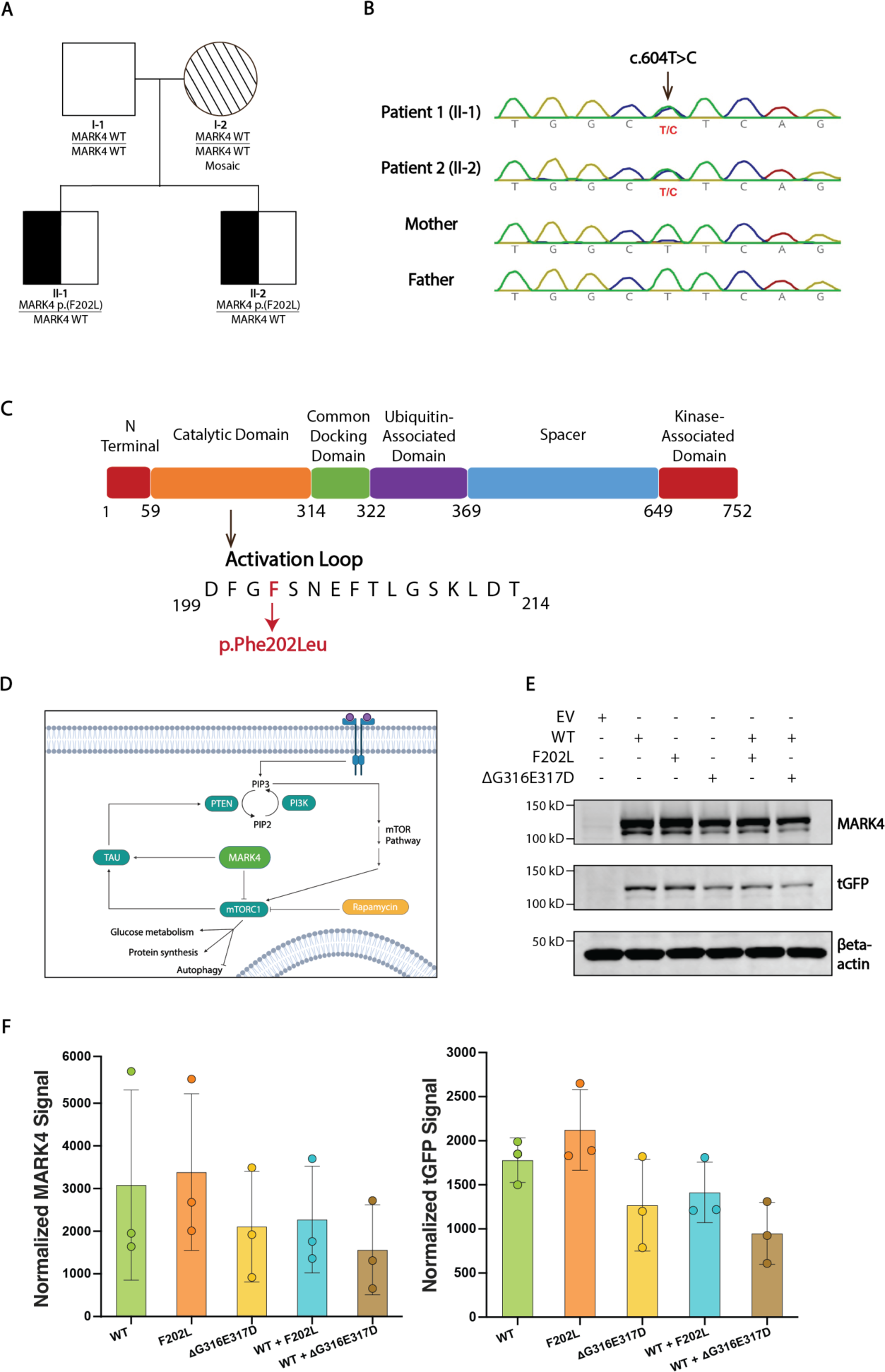
Clinical phenotype of siblings with novel *MARK4* variant, neurodevelopmental presentation, and dysmorphic features. **(A)** Family pedigree. Half-filled symbols=heterozygous affected individuals (Patient 1 and Patient 2), Lined symbol=unaffected, mosaic individual. **(B)** Sanger sequencing of DNA extracted from whole blood of Patient 1, Patient 2, and their parents. Site of substitution is indicated. **(C)** Schematic illustrating the protein domains of MARK4. Location of variant in the activation loop shown in red. **(D)** Schematic illustrating the link between MARK4, tau, and the mTOR pathway. **(E)** Representative immunoblot of lysates obtained from HEK293 cells transfected with plasmids expressing tGFP-tagged MARK4^wt^, the p.F202L variant, the p.ΔG316E317D variant, and/or an EV control. The amount of MARK4 and tGFP was monitored with anti-MARK4 and anti-tGFP antibodies. Equal loading was confirmed with an anti-βeta-actin antibody. Uncropped immunoblot can be found in Supplementary Fig. 1. **(F)** Band intensities were quantified and normalized signals for MARK4 and tGFP were obtained using Empiria Studio® Software. Data from three independent experiments is shown (means ± sd). One-way ANOVA followed by a Dunnett post-hoc test for multiple comparisons.

### MARK4 variant localizes to the activation loop of the kinase

The canonical sequence of MARK4 is 752 amino acids long and is split into six domains (Fig. 1C).^13^ The p.F202L variant localizes to the activation loop of MARK4 inside the catalytic domain. A highly-conserved aspartate-phenylalanine-glycine (DFG) motif initiates the activation loop. The DFG motif can switch between two conformations and alter the accessibility of the ATP binding site and thus activate or inactive the kinase.^13^ The p.F202L variant causes an amino acid substitution adjacent to the DFG motif. Based on its position, the variant was predicted to impact kinase activity of MARK4 thus affecting the phosphorylation of downstream targets (Fig. 1D).

### The p.F202L variant does not alter MARK4 expression

We quantified the impact of the p.F202L variant on MARK4 protein expression by immunoblotting. We used HEK293 cells as a model to study MARK4 expression and activity because they have low endogenous expression of MARK4, lack tau expression, and have previously been used to study MARK4 biology.^9,10^ We included the p.ΔG316E317D variant in our experiments as a control. Cells were transfected with plasmids expressing WT and variant MARK4 proteins or an empty vector (EV) control. A heterozygous state was modelled by co-transfecting cells with WT and variant MARK4. A prominent band at approximately 110 kDa was detected by MARK4 and tGFP antibodies in all wells except those transfected with the EV (Fig. 1E and Supplementary Fig. 1). The p.ΔG316E317D variant had no impact on protein expression, replicating previous research findings (Fig. 1F).^9,10^ The p.F202L variant also had no impact on MARK4 and tGFP expression.

### The p.F202L variant causes increased tau phosphorylation

In the adult brain, six tau isoforms are expressed from a single *MAPT* gene by alternative splicing of exons 2, 3, and 10.^14^ 0N3R tau is the most abundant in the fetal brain and lacks exons 2, 3, and 10, making it the smallest isoform.^14^ 2N4R tau retains exons 2, 3, and 10, making it the largest isoform in adult brains.^14^

Since the patients presented with childhood-onset neurological symptoms, we first investigated the impact of the p.F202L variant on 0N3R tau phosphorylation. We performed phospho-flow cytometry on cells co-transfected with 0N3R tau and MARK4 proteins (see Supplementary Fig. 2A for the gating strategy). Transfection with the p.F202L variant resulted in almost double the amount of phospho-tau compared to transfection with MARK4^wt^ (63.5% vs. 36.4% median phospho-tau, p < 0.0001) (Fig. 2A and B). The p.ΔG316E317D variant,^9,10^ as expected, resulted in increased phospho-tau compared to MARK4^wt^ (49.5% vs. 34% median phospho-tau, p < 0.05). Notably, the p.F202L variant resulted in increased phospho-tau compared to the p.ΔG316E317D variant (63.5% vs. 49.5% median phospho-tau, p < 0.01). Cells that were co-transfected with the biochemically null MARK4 p.D199A variant^7^ or an EV control had very low levels of tau phosphorylation (4.3% and 8.6% median phospho-tau). Similar results were observed with the 2N4R tau isoform as the 0N3R tau isoform (Fig. 2C and D). Thus, the p.F202L MARK4 variant causes increased phosphorylation of tau isoforms found in the fetal and adult brain.

**Figure 2.**
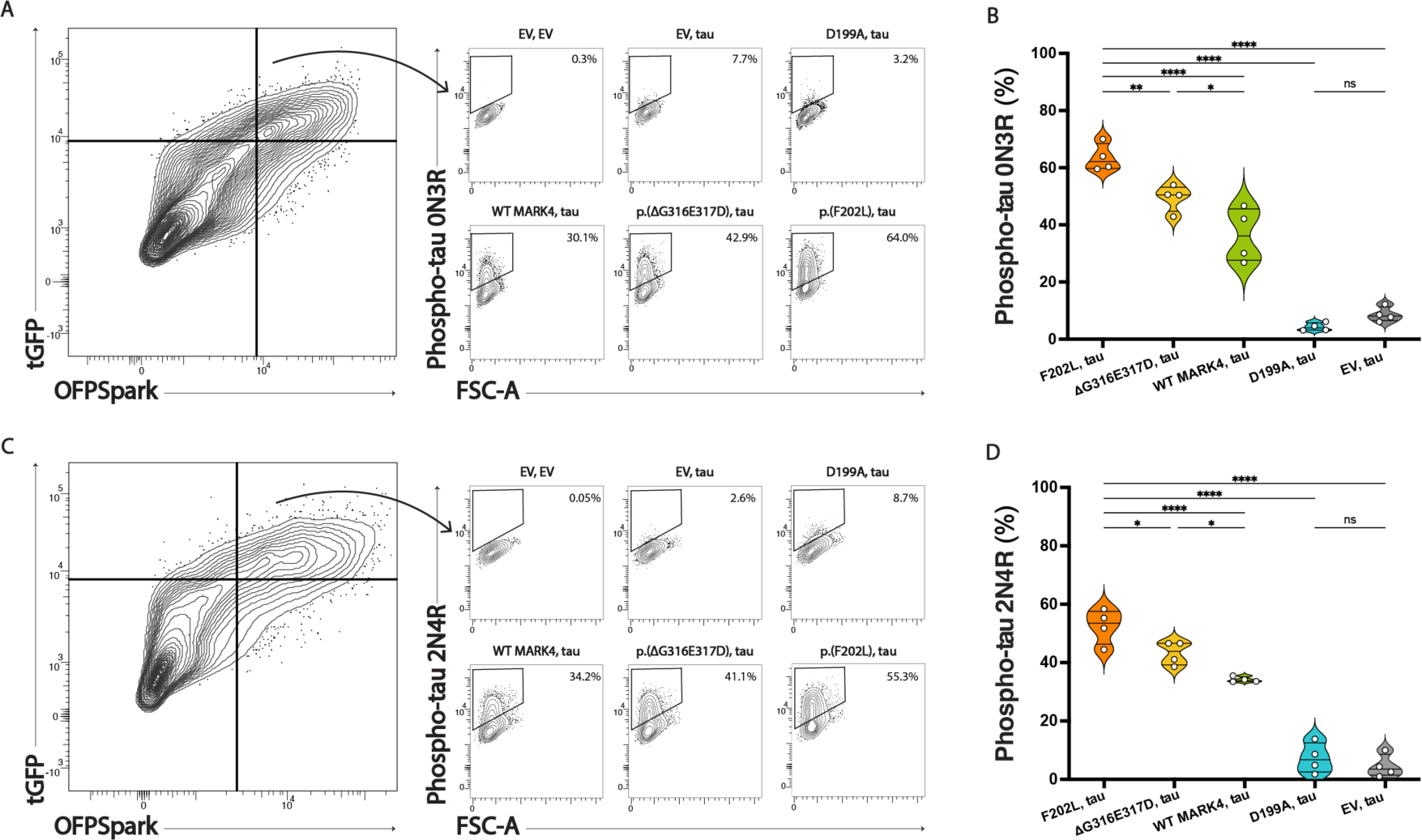
The p.F202L variant causes increased tau phosphorylation in the fetal and adult brain. **(A)** Contour plots showing phospho-tau S262 in cells co-transfected with 0N3R tau and MARK4 proteins. Gating strategy to identify tGFP^+^OFPSpark^+^ cells can be found in Supplementary Fig. 2a. **(B)** Violin plot showing percentage of cells with phosphorylated 0N3R tau; median and interquartile interval are indicated by black bars (N=4). **(C)** Contour plots showing phospho-tau S262 in cells co-transfected with 2N4R tau and MARK4 proteins. **(D)** Violin plot showing percentage of cells with phosphorylated 2N4R tau; median and interquartile interval are indicated by black bars (N=4). One-way ANOVA followed by a Dunnett post-hoc test for multiple comparisons. \**P* < 0.05; \*\**P* < 0.01; \*\*\*\**P* < 0.0001. FSC: forward scatter.

### The p.F202L variant causes upregulation of the mTORC1 pathway

Since MARK4 is a known negative regulator of mTORC1,^7^ we next analyzed the effect of the p.F202L variant on the kinase’s ability to inhibit the mTORC1 pathway. We studied this by quantifying the phosphorylation of ribosomal protein S6, a well-established downstream marker of mTORC1 pathway activation.^15^ Cells were transfected with WT and variant MARK4 proteins or an EV control to determine their impact on S6 phosphorylation, which exists in a phosphorylated state at baseline in HEK293 cells. Fluorescence minus one (FMO) control was used for identifying cells with phospho-S6 (S235/S236) (see Supplementary Fig. 2B for the gating strategy). Flow cytometric analysis of transfected (tGFP^+^) cells revealed that the p.F202L variant, when compared to MARK4^wt^, had increased levels of phospho-S6 and thus upregulation of the mTORC1 pathway (45.7% vs. 29.1% median phospho-S6, p < 0.01) (Figure 3A and B). In this assay, the p.F202L and p.ΔG316E317D variants had similar levels of phospho-S6 (45.7% and 44.9% median phospho-S6). As a negative control, flow cytometric analysis of tGFP^-^ cells revealed comparable levels of phosphorylated S6 in all conditions (Fig. 3A and C), confirming that expression of the *MARK4* variants was required to generate differences in the level of phosphorylated S6 protein.

**Figure 3.**
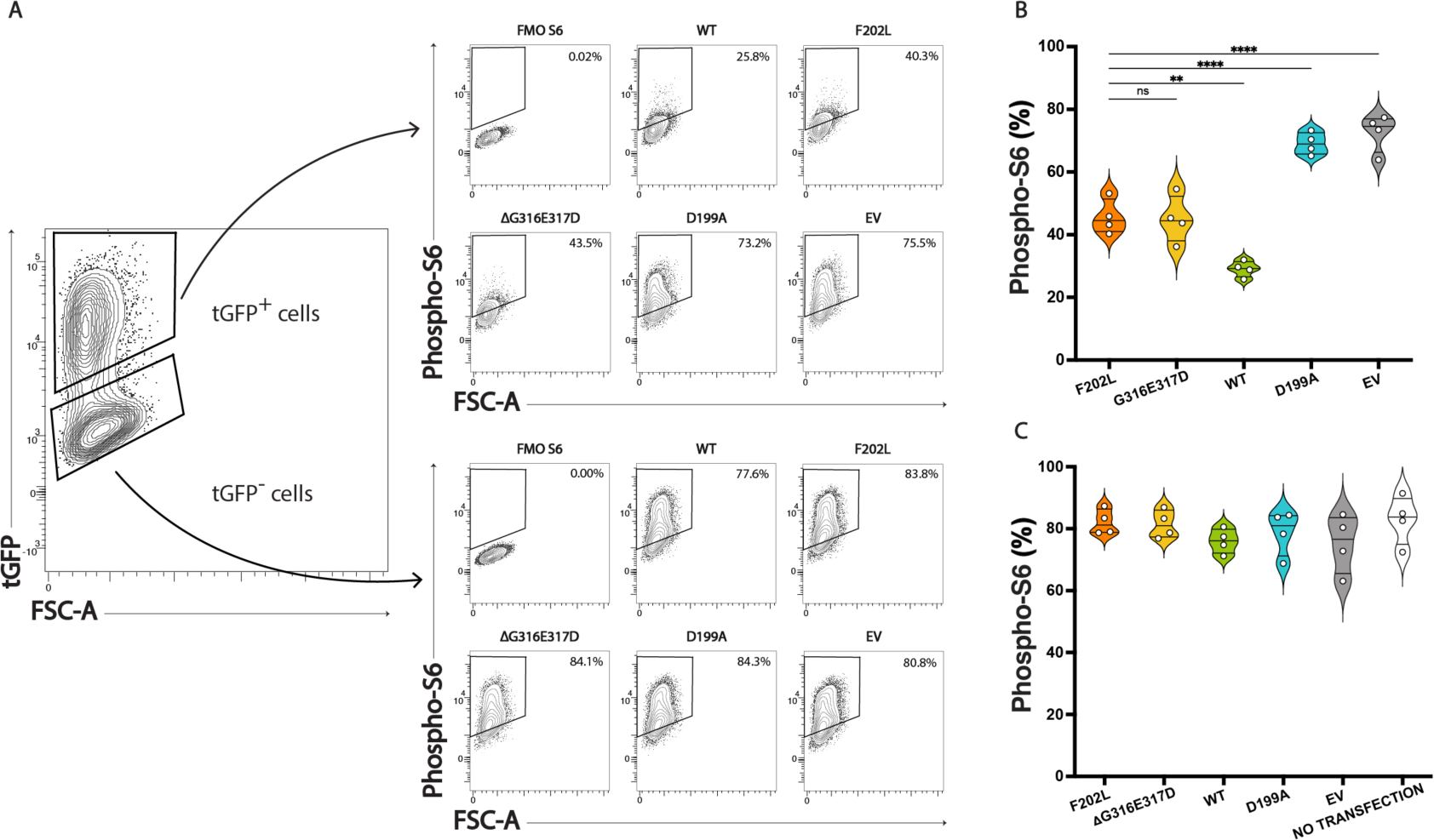
The p.F202L variant results in increased phosphorylation of S6 and upregulation of the mTOR pathway. **(A)** Contour plots showing phospho-S6 S235/S236 in cells transfected with MARK4. Gating strategy to identify tGFP^+^ cells can be found in Supplementary Fig. 2b. **(B)** Violin plot showing percentage of tGFP^+^ cells with phosphorylated S6; median and interquartile interval are indicated by black bars (N=4). **(C)** Violin plot showing percentage of tGFP^-^ cells with phosphorylated S6; median and interquartile interval are indicated by black bars (N=4). One-way ANOVA followed by a Dunnett post-hoc test for multiple comparisons. \*\**P* < 0.01; \*\*\*\**P* < 0.0001. FSC: forward scatter.

### The p.F202L variant causes increased tau phosphorylation independent of mTORC1 activity

We sought to determine if the p.F202L variant causes increased phospho-tau directly through MARK4 interaction or indirectly through mTORC1 activity by treating cells with rapamycin, a known mTORC1 inhibitor (Fig. 1D).^16^ We measured the impact of rapamycin on tau phosphorylation in cells co-transfected with 0N3R tau and MARK4 proteins. Flow cytometric analysis of double-positive cells (GFP^+^ and OFPSpark^+^) revealed no differences in phospho-tau in cells in which rapamycin was present or absent for MARK4^wt^, the p.F202L variant, and the p.D199A variant (Fig. 4A and Supplementary Fig. 3A). Next, to document rapamycin’s biological activity, we measured the impact of rapamycin on phospho-S6. As expected, flow cytometric analysis of tGFP^+^ revealed decreased S6 phosphorylation in cells treated with rapamycin compared to cells lacking rapamycin (Fig. 4B and Supplementary Fig. 3B). Taken together, these results indicate that the p.F202L variant causes a gain-of-function in the ability of MARK4 to phosphorylate tau, independent of the impact that MARK4 has on the mTORC1 pathway.

**Figure 4.**
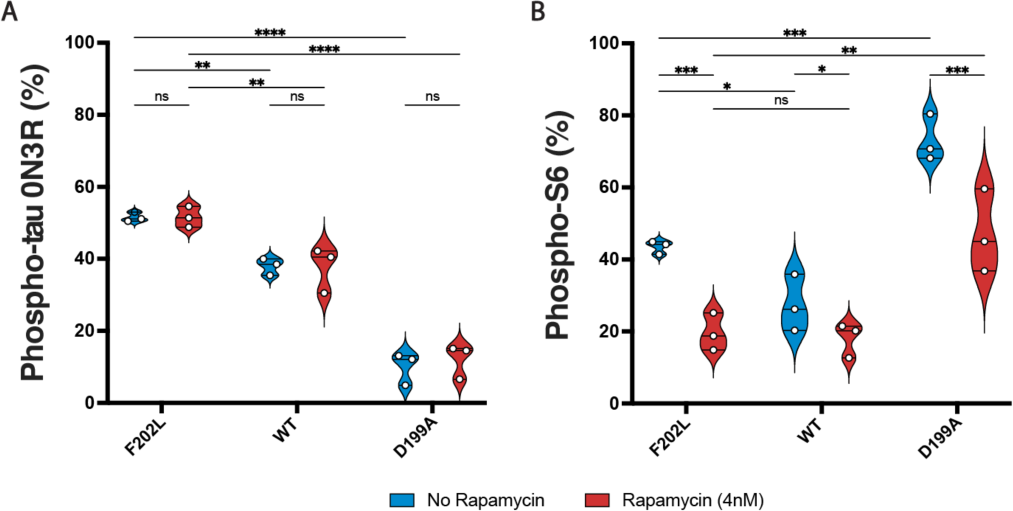
The p.F202L variant causes a gain-of-function in the ability of MARK4 to phosphorylate tau. **(A)** Violin plot showing percentage of tGFP^+^OFPSpark^+^ cells with phospho-tau in control and rapamycin-treated cells expressing MARK4^wt^, the p.F202L variant, and the p.D199A variant; median and interquartile interval are indicated by black bars (N=3). Individual contour plots can be found in Supplementary Fig. 3a. **(B)** Violin plot showing percentage of tGFP^+^ cells with phospho-S6 in control and rapamycin-treated cells expressing MARK4^wt^, the p.F202L variant, and the p.D199A variant; median and interquartile interval are indicated by black bars (N=3). Individual contour plots can be found in Supplementary 3b. Two-way ANOVA with Tukey’s multiple comparison test. \**P* < 0.05; \*\**P* < 0.01; \*\*\**P* < 0.001; \*\*\*\**P* < 0.0001.

## Discussion

Here we present clinical, genetic, and functional evidence of a potential new neurodevelopmental disorder caused by a heterozygous p.F202L gain-of-function variant in MARK4. The variant is maternally inherited from the patients’ unaffected mother who has low level mosaicism (11 of 97 reads were variant in blood derived DNA). The variant is located in the activation loop of MARK4 and has no effect on protein expression, eliminating haploinsufficiency as the disease mechanism (Fig. 1E). Functional investigation revealed that the variant results in a gain-of-function in the ability of MARK4 to phosphorylate tau and leads to upregulation of the mTORC1 pathway (Fig. 2-3). Additional cases harboring *MARK4* gain-of-function variants will need to be identified to define the full spectrum of this new disorder. Identification of additional cases will establish if there is a consistent or recognizable set of morphological differences caused by gain-of-function variants in MARK4; in these siblings, consistent features included hypertelorism (HP:0000316), ptosis (HP:0000508), a shortened columella (HP:0002000) with coarseness to the alae nasi (HP:0009928), pectus excavatum (HP:0000767), and prominent fetal pads (HP:0001212).

The role of tau in normal cognitive aging is well established. In tauopathies, however, tau becomes abnormally hyperphosphorylated and undergoes pathological changes; it disassociates from microtubules, self-aggregates to oligomers, and forms paired helical filaments (PHFs), and neurofibrillary tangles.^1-3^ In addition to its role in pathological aging, tau also plays a critical role in early development, facilitating neuronal differentiation and migration, synaptogenesis, and dendritic spine formation.^4,5^ Alteration of these processes leads to neurodevelopmental disorders.^17-19^

We propose that increased MARK4-mediated phosphorylation of tau at S262 impairs tau’s normal functions and causes the childhood-onset neurological presentation and dysmorphic features in Patients 1 and 2. Located in the microtubule-binding domain of tau, S262 plays a critical role in regulating microtubule binding and forming PHF-tau in Alzheimer’s disease.^20,21^ Notably, both patients present with a similar phenotype to individuals with haploinsufficiency of *MAPT*, the gene encoding tau.^22^ Developmental delay, intellectual disability, hypotonia, and dysmorphic craniofacial features are consistently reported in *MAPT*^+/-^ individuals. Seizures and abnormal hearing loss are reported in some *MAPT*^+/-^ individuals. The parallels in clinical presentation between Patients 1 and 2 and *MAPT*^+/-^ individuals suggest that increased tau phosphorylation at S262 has similar downstream consequences as lacking a copy of the *MAPT* gene. Since phosphorylation of S262 is developmentally regulated,^23^ persistent phosphorylation at this residue in the developing brain may compromise protein function, priming tau for further phosphorylation by other kinases, and ultimately generating pathological tau.

In cells, protein aggregates can be eliminated by autophagy. Autophagy is regulated by the mTOR pathway, which also plays an extensive role in signaling pathways centered on energy availability, stimulation of anabolic molecular processes, and cell growth (Fig. 1D).^24^ Abnormal activation of the mTOR pathway is implicated in many human diseases. Reminiscent of our finding with the MARK4 p.F202L variant, upregulation of the mTOR pathway along with hyperphosphorylated tau is observed in several neurodevelopmental disorders, including hemimegalencephaly, tuberous sclerosis complex, and focal cortical dysplasia.^5,25^ Furthermore, the mTOR pathway is upregulated in mouse models of autism and Alzheimer’s disease.^26^ Increased activity of the mTOR pathway suppresses autophagy in cells^27^; thus, continued suppression or dysfunction of autophagy prevents removal of protein aggregates, and is linked to many neurodegenerative disorders.^28^ Suppression of autophagy may also be contributing to our patients’ phenotype since the p.F202L variant results in upregulation of the mTOR pathway. Future studies might examine the therapeutic potential of rapamycin or other mTOR inhibitors in patients with gain-of-function variants in *MARK4*.

The MARK family of proteins is increasingly recognized to play an important role in the developing brain, and *MARK2* has been identified as a risk gene for autism.^29,30^ Here we expand the role of MARK proteins in neurodevelopment by showing that a monoallelic gain-of-function *MARK4* variant associates with the childhood-onset phenotype of developmental delay, intellectual disability, and dysmorphism. With the recent development and approval of therapeutics that target tau aggregation, the role that these may play in modifying the natural history of this and other neurodevelopmental disorders associated with *MARK4* gain-of-function should be explored.

## Data Availability

Data supporting the findings of this study are available from the corresponding author, upon reasonable request.

## Acknowledgements

We thank the patients and their parents for participation in this study. We further thank the Advisory Committee of the Rare Disease Discovery Hub.

## Abbreviations

DFG: aspartate-phenylalanine-glycine
EV: Empty vector
FMO: Fluorescence minus one
PHF: paired helical filaments
tGFP: Turbo green fluorescent protein
WT: Wild-type

