## Supplementary material for "Gain-of-function MARK4 variant associates with pediatric neurodevelopmental disorder and dysmorphism"

#### Contents

|  | **Title** | **Page** |
| --- | --- | --- |
| Supplementary Methods | Materials and methods | 2-4 |
| Case Reports | Case Reports | 5-6 |
| Supplementary Tables | Supplementary Table 1: Sequence of oligonucleotides | 7 |
|  | Supplementary Table 2: Clinical summary of patients with the p.(F202L) MARK4 variant | 8 |
|  | Supplementary Table 3: Variant description of Patients 1 and 2 | 9 |
| Supplementary Figures | Supplementary Figure 1 Full-length immunoblot shown in Fig. 1E | 10 |
|  | Supplementary Figure 2 Gating strategy for phospho-flow experiments | 11 |
|  | Supplementary Figure 3 Contour plots for rapamycin experiments | 12 |
| Supplementary References | References | 13 |

### Materials and methods

#### Exome sequencing, analysis and variant validation

Trio-based whole exome sequencing was performed on genomic DNA from Patient 1 and both parents on an Illumina platform at Ambry Genetics after capture of targeted regions using xGen™ Exome Hybridization Panel (Cat# 10005152; IDT).^1^ More than 99% of targeted regions were covered by at least ten reads that averaged 150 bp in length. Reads were aligned to GRCh37/Hg19 and variants were called using BWA-MEM, GATK 3.5-0, SAMtools 0.1.19, and Picard 1.139, and then annotated and prioritized with VarSeq v.1.5 (Golden Helix).

The *MARK4* variant was confirmed by Sanger sequencing of genomic DNA from the patients and their parents. Genomic DNA was extracted from whole blood using a QIAmp DNA Blood Mini Kit (Cat# 51106; Qiagen) according to the manufacturer’s recommendations. The variant region was amplified by PCR using GoTaq Green Master Mix (Cat# M7122; Promega) and primers that flank the area (Supplementary Table 2). The PCR product was purified using a QIAquick PCR Purification Kit (Cat# 28106; Qiagen) and sequenced using the same primers.

#### Generation and expression of MARK4 and tau proteins

Plasmids used for transfection studies contained full-length *MARK4* cDNA in a pCMV6-AC-GFP vector (Cat# RG234829; OriGene Technologies) and *MAPT* (tau) 0N3R cDNA in a pCMV3-OFPSpark vector (Cat# HG10058-ACR; Sino Biological). Appropriate empty vectors, pCMV6-AC-GFP (Cat# PS100010; OriGene Technologies) and pCMV3-OFPSpark (Cat# CV025; Sino Biological) were also used for transfection studies.

To generate *MARK4* variants, c.604T>C, c.946_951delGGTGAGinsGAT, and c.596A>C, a Q5 Site-Directed Mutagenesis Kit (Cat# E0554S; New England Biolabs) was used according to the manufacturer’s recommendations, with primer pairs noted in Supplementary Table 1. A plasmid expressing the 2N4R isoform of tau was generated using the tau 0N3R OFPSpark plasmid. A BamHI restriction site was introduced between the tau 0N3R sequence and the C-terminal OFPSpark tag, using a Q5 Site-Directed Mutagenesis Kit and primer pair listed in Supplementary Table 1. The cDNA sequence for the 2N4R isoform was purchased from GenScript and cloned into the OFPSpark vector simultaneously as the cDNA sequence for tau 0N3R was cloned out. Cloning was accomplished using the following restriction enzymes, KpnI-HF (Cat# R3142S; New England Biolabs) and BamHI-HF (Cat# R3136S; New England Biolabs). All variant plasmids were purified from 10-beta competent *E. coli* using a QIAprep Spin Miniprep Kit (Cat# 27106; Qiagen) and confirmed by Oxford Nanopore technology (Plasmidsaurus).

Transient expression of the plasmids in HEK293 cells was accomplished using a Lipofectamine 3000 Kit (Cat# L3000015; Thermo Fisher Scientific) according to the manufacturer’s recommendations. For immunoblotting experiments, 1.0 × 10^6^ HEK293 cells were seeded in 6-well culture plates in 2 mL of Dulbecco modified Eagle medium (DMEM; Cat# SH3002201; GE Healthcare Hyclone) supplemented with 10% FBS (Cat# A3840302; Gibco, Life Technologies) and 1mM sodium pyruvate (Cat# 11360070; Gibco, Life Technologies). Cells were incubated at 37°C for 24 hours and then transfected with 3 µg of plasmid DNA using the Lipofectamine^TM^ 3000 Transfection Reagent and harvested after 24 hours. For flow cytometry experiments, 2.0 × 10^5^ HEK293 cells were seeded in 24-well culture plates in 0.5 mL of DMEM supplemented with 10% FBS and 1mM sodium pyruvate. Cells were incubated at 37°C for 24 hours and then transfected with 500 ng of plasmid DNA and harvested after 24 hours.

#### Immunoblotting

Expression of wild-type MARK4 and variants was determined by immunoblotting. Cells were harvested in chilled radioimmunoprecipitation assay buffer (Cat# 89901; Thermo Fisher Scientific) supplemented with HALT protease and phosphatase inhibitor cocktail (Cat# 87786; Thermo Fisher Scientific) and then lysed for 15 minutes on ice. Cell lysates (15 µg – within linear range of detection for all antibodies) were separated by 10% SDS-polyacrylamide gel electrophoresis and transferred onto polyvinylidene difluoride membranes (Cat# IPFL00010; Immobilon-FL; MilliporeSigma). Membranes were blocked with 5% BSA in TRIS-buffered saline with Tween-20 for an hour, and then incubated with primary antibodies in blocking buffer overnight at 4˚C. The next day, membranes were washed and incubated with secondary antibodies for 1 hour at room temperature and then imaged using a LI-COR Odyssey infrared scanner (LI-COR Biosciences). Three primary antibodies were used in these experiments: MARK4 (Cat# 4834S; Dilution: 1:500; Cell Signaling Technologies), tGFP (Cat# TA150041; Dilution: 1:500; OriGene Technologies), and βeta-Actin (Cat# 3700S; Dilution: 1:40,000; Cell Signaling Technologies). The following two secondary antibodies were used to detect the target proteins, goat anti-rabbit IgG DyLight 800 conjugated (Cat# 611-145-002-0.5; Dilution: 1:20,000; Rockland Immunochemicals) and goat anti-mouse IgG IRDye 680RD (Cat# 926-6870; Dilution: 1:20,000; LI-COR).

#### Flow cytometry

Phosphorylation of S6 and tau proteins was quantified in transfected HEK293 cells using phospho-flow cytometry. Harvested cells were washed with PBS and stained with a Fixable Viability Stain (Cat# 565388; Dilution: 1:100; BD Biosciences) for 5 minutes at room temperature. The viability stain was removed with a PBS wash, and then cells were fixed with Cytofix (Cat# 554655; BD Biosciences) for 10 minutes at room temperature and permeabilized with Phosflow PermBuffer III (Cat# 558050; BD Biosciences) for 15 minutes at 4°C. Permeabilized cells were washed with Perm Wash Buffer (Cat# 554723; BD Biosciences) and stained with intracellular flow antibodies. Phosphorylated S6 was detected by staining with an anti-S6 phospho (Ser235/Ser236) antibody (Cat# 608604; Dilution: 1:25; BioLegend) in the dark for 30 minutes at room temperature. Phosphorylated tau was detected by staining with an anti-tau phospho (Ser262) antibody (Cat# 44-750G; Dilution: 1:25; Thermo Fisher Scientific) for 2 hours at room temperature followed by staining with a goat anti-rabbit secondary antibody (Cat# A-31556; Dilution: 1:25; Thermo Fisher Scientific) in the dark for 30 minutes at room temperature. Stained samples were acquired on a BD LSRFortessa™ Cell Analyzer and the generated data were analyzed using FlowJo software (BD Biosciences).

We determined that the MARK4 variant causes a gain-of-function in the ability of MARK4 to phosphorylate tau using rapamycin (Cat# AY-22989; MedChemExpress). Rapamycin (4nM) was added to transfected cells after 6 hours. Twenty-four hours after transfection, cells were prepared for phospho-flow cytometry as described above.

### Case Reports

#### Patient 1

Patient 1 is a male (ages 17-22 years) born to healthy, non-consanguineous parents. (Supplementary Table 1). His younger brother has similar phenotypic findings and is Patient 2 of this study. There is no additional significant family history. Patient’s mother had an unremarkable pregnancy and he was born at term following a prolonged labor which required vacuum extraction. At birth he had a weight of 2.9 kg and required intubation with mechanical ventilation. Early feeding challenges and weight loss caused him to be readmitted to the hospital and fed through a nasogastric tube. He presented with hypotonia and recurrent hypoglycemia during the neonatal stage.

All early developmental milestones were delayed. He had early failure-to-thrive. His body mass index (BMI) has been persistently lower than the 3^rd^ percentile since childhood. When he began walking, he was initially somewhat uncoordinated and had difficulty with some gross and fine motor movements. As a young child (ages 3-8 years), he was diagnosed with expressive language delay and began to have daily generalized tonic-clonic seizures. Several medications (carbamazepine, valproate, and topiramate) were tried to manage his seizures until he responded well to daily, combined treatment with lamotrigine and levetiracetam. As a child, he had difficulties with his behaviors, attention span, hyperactivity, and impulsivity, culminating in a diagnosis of an attention deficit hyperactivity disorder (ADHD). Treatment with amphetamine/dextroamphetamine salts was discontinued due to paradoxical worsening of behaviors as well as decreased sleep and appetite, he was subsequently started on atomoxetine. He was also noted to have social difficulties, limited imaginative play, and sensory sensitives. He underwent assessments for autism, cognitive functioning (WPPSI), and adaptive behavior skills (Vineland). He was diagnosed with autism and had low cognitive skills (1^st^ percentile) and moderately low adaptive skills (3^rd^ percentile). He required a full-time aide at school because of his moderate-to-severe intellectual disability.

He has dysmorphic features, causing him to resemble his brother but not their parents. The facial dysmorphism includes prominent supraorbital ridges and eyebrows, hypertelorism with an interpupillary distance of 6.6 cm, and downward-slanting palpebral fissures. Additionally, he has a depressed nasal bridge and a prominent bulbous appearance to his nose, with coarseness to his alae nasi and a shortened columella. Facial asymmetry was also noted, with the left ear being slightly lower than the right ear, although they are normal in size at 6.4 cm. He has a prominent mandible and his lips have a “tent-shaped” appearance with coarseness. Examination of the chest revealed translucent skin and a mild pectus excavatum, slightly asymmetrical with the right side greater than the left side. Examination of the hands showed symmetrical brachydactyly and prominence of fetal pads on most of his digits. Increased carrying angle is observed at his elbows. Neurobiological examination revealed finger-to-nose testing was slightly abnormal with a mild tremor bilaterally. He was noted to have difficulties completing a tandem Romberg test.

Extended metabolic work-up and a brain MRI (ages 3-8 years) were both normal. In terms of his initial genetic work-up, chromosomal microarray and multi-gene panels for both intellectual disability/autism and glycogen storage diseases were non-diagnostic.

#### Patient 2

Patient 2 is also a male (ages 11-16 years) and the younger brother of Patient 1. He was born at term following an unremarkable pregnancy. He presented with a mild learning disability and behavioral challenges. As a young child (ages 3-8 years), his mother became concerned regarding early learning, speech, and language development. During childhood, he was diagnosed with ADHD and had frequent ear infections. He has abnormal hearing loss related to cholesteatoma development, and thus uses hearing aids. He has never had seizures or other medical concerns. Measurements for his BMI were only available at two timepoints; at one of the two timepoints, his BMI was below the 3^rd^ percentile.

Similar to his older brother, he has dysmorphic features, causing the brothers to have a distinctive appearance. The facial dysmorphism includes hypertelorism with an interpupillary distance of 6.5 cm, ptosis, and a prominent bulbous appearance to his nose, with coarseness to his alae nasi and a shortened columella. Examination of the chest revealed a mild pectus excavatum with asymmetry to the left. Prominence of fetal pads was observed on all of his digits. Examination of his extremities showed increased carrying angle at the elbows bilaterally. Neurobiological examination revealed no obvious cerebellar findings. Finger-to-nose testing was normal and he did not have difficulties completing a tandem Romberg test. Chromosomal microarray testing was done to investigate the cause of his intellectual disability but was determined to be non-diagnostic.

**Supplementary Table 1: Sequence of oligonucleotides**

| ***MARK4* primers for variant validation in both patients and parents** | | |
| --- | --- | --- |
| Template | Direction | Sequence (5' - 3') |
| genomic DNA (blood) | Forward | TCATAATTCTCAAACTACGCTATGAAA |
| genomic DNA (blood) | Reverse | GATAGGGGCCTGACTGTGG |
| **Site-directed mutagenesis primers for MARK4 p.F202L variant** | | |
| Template | Direction | Sequence (5' - 3') |
| Wild-type MARK4 plasmid | Forward | TGACTTTGGCCTCAGCAACGA |
| Wild-type MARK4 plasmid | Reverse | GCAATCTTGATGTTGGCC |
| **Site-directed mutagenesis primers for MARK4 p.ΔG316E317D variant** | | |
| Template | Direction | Sequence (5' - 3') |
| Wild-type MARK4 plasmid | Forward | GATGAGTTGAAGCCATACACAG |
| Wild-type MARK4 plasmid | Reverse | CTCATAGCCGATGTTGATC |
| **Site-directed mutagenesis primers for MARK4 p.D199A variant** | | |
| Template | Direction | Sequence (5' - 3') |
| Wild-type MARK4 plasmid | Forward | AAGATTGCTGCCTTTGGCTTCAGC |
| Wild-type MARK4 plasmid | Reverse | GATGTTGGCCTCGGCATC |

**Supplementary Table 2: Clinical summary of patients with the MARK4 p.F202L variant**

|  | **Patient 1** | **Patient 2** |
| --- | --- | --- |
| Parental consanguinity | No | |
| Family history | Non-contributory | |
| Sex | Male | |
| **Pregnancy and Birth** | | |
| Antenatal complications | Uncomplicated | Uncomplicated |
| Neonatal findings | Hypotonia and hypoglycemia, stayed in NICU for one week | Normal neonatal stage |
| **Manifestation** | | |
| First clinical signs | Recurrent hypotonia and hypoglycemia | Delayed early learning, speech and language development |
| **Most Recent Examination** | | |
| Age (years) | 17-22 | 11-16 |
| Weight, kg/z-score | 45.8/-3.19 | 44.8/+0.39 |
| Height, cm/z-score | 171/-0.76 | 165.4/+1.79 |
| Head circumference, cm/z-score | 56/-0.44 | 53.4/-0.72 |
| Neurobiological examination | Slightly abnormal finger-to-nose testing with a mild tremor bilaterally; difficulties performing tandem Romberg | Normal finger-to-nose testing; slight tremor; no difficulties completing a tandem Romberg test |
| **Neurobiological Features** | | |
| Global developmental delay | Slow gain in developmental milestones | Slow gain in developmental milestones |
| Intellectual disability | Moderate-to-severe intellectual disability | Moderate intellectual disability |
| Behaviour | ADHD; behavioural abnormalities | ADHD; behavioural abnormalities |
| Language and speech | Delayed speech development; diagnosed with expressive language delay | Delayed speech development |
| Seizures | Daily generalized tonic-clonic seizures; seizures are now well-managed by 2 medications | No concerns |
| MRI scan | Normal | Not performed |
| **Other Findings** | | |
| Hearing | No concerns | Abnormal hearing loss; has hearing aids |
| Feeding | Feeding difficulties after birth; unable to be breastfed and thus fed through a tube | Some feeding difficulties during childhood |
| Dysmorphic features | Hypertelorism, mild ptosis, shortened columella with coarseness to the alae nasi and prominence to the nasal tip, facial asymmetry, mild pectus excavatum, prominent fetal pads, increased carrying angle at the elbows, and translucent skin | Hypertelorism, mild ptosis, shortened columella with coarseness to the alae nasi and prominence to the nasal tip, mild pectus excavatum, prominent fetal pads, and increased carrying angle at the elbows |

ADHD = attention deficit hyperactivity disorder; NICU = neonatal intensive care unit.

**Supplementary Table 3: Variant description of proband and younger brother**

| Chromosome | 19 |
| --- | --- |
| Genomic position (GRCh38) | 45,271,526 |
| cDNA position (NM_001199867.1) | c.604T>C |
| Nucleotide reference | T |
| Protein variant (NP_001186796.1) | p.(Phe202Leu) |
| Exon affected | 8 |
| Zygosity | Heterozygous |
| Inheritance | Maternally inherited |
| CADD (v1.6) | 27 |
| REVEL | 0.313 |

CADD = combined annotation dependent depletion; REVEL = Rare exome variant ensemble learner.

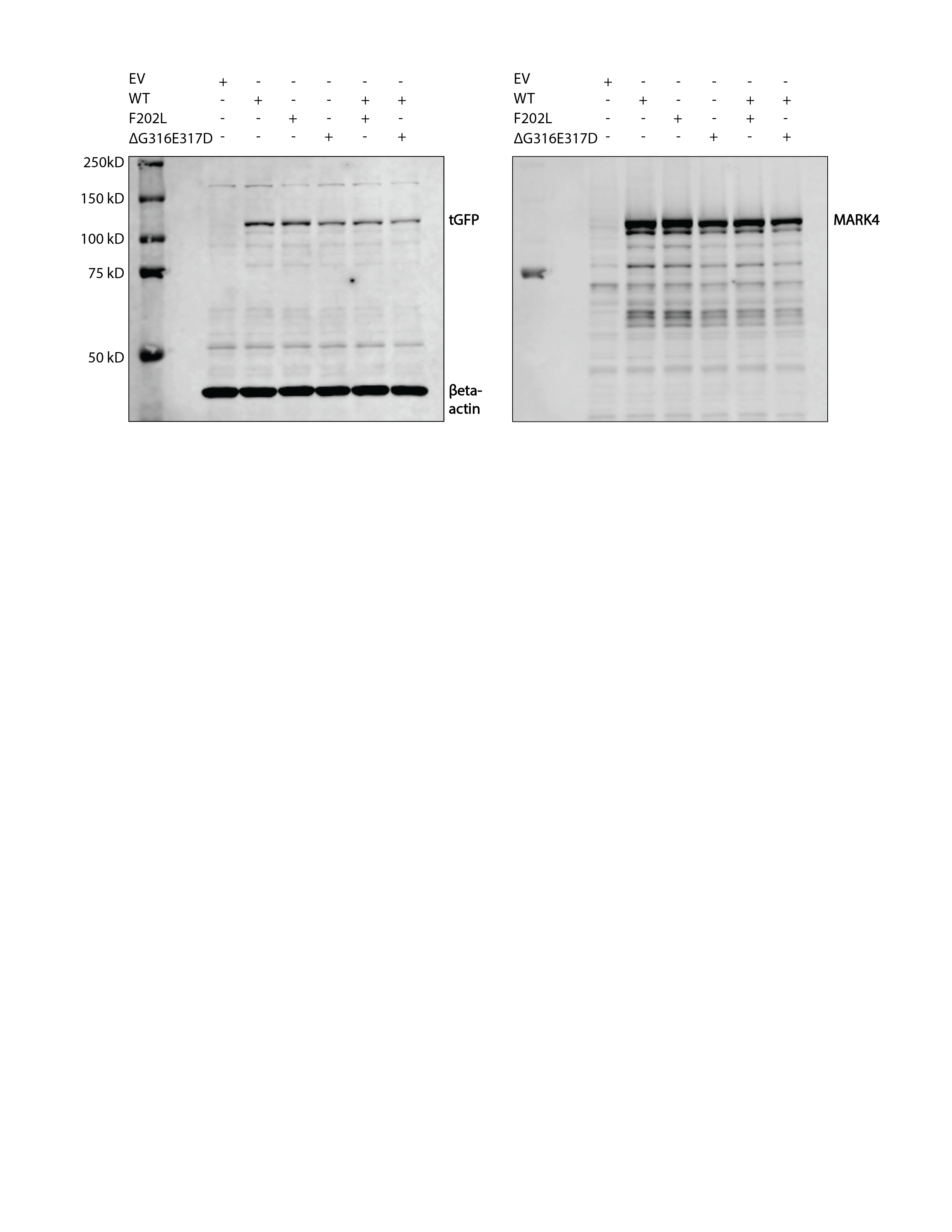

**Supplementary Figure 1 Full-length immunoblot shown in Fig. 1E.** Full-length immunoblot of lysates obtained from HEK293 cells transfected with plasmids expressing tGFP-tagged MARK4^wt^, the p.F202L variant, the p.ΔG316E317D variant, and/or an empty vector (EV) control. The immunoblot was probed with antibodies against MARK4, tGFP, and βeta-actin. Equal loading was demonstrated with the anti-βeta-actin antibody. Quantification of band intensities for MARK4, tGFP, and βeta-actin fluorescence signals was done using Empiria Studio® Software.

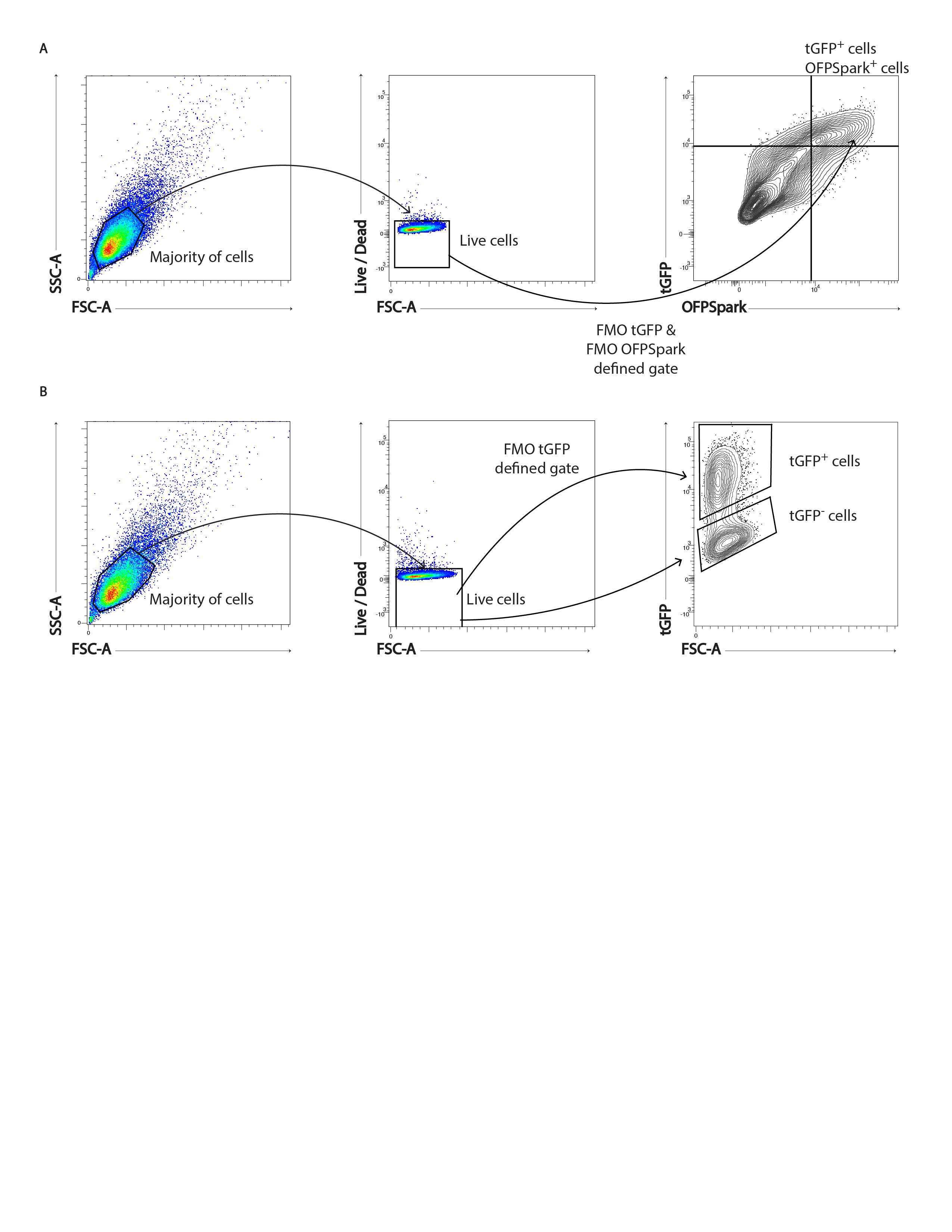

**Supplementary Figure 2 Gating strategies for phospho-flow experiments. (A)** Gating strategy for phospho-tau flow experiments to identify tGFP^+^OFPSpark^+^ cells. **(B)** Gating strategy for phospho-S6 flow experiments to identify tGFP^+^ cells.

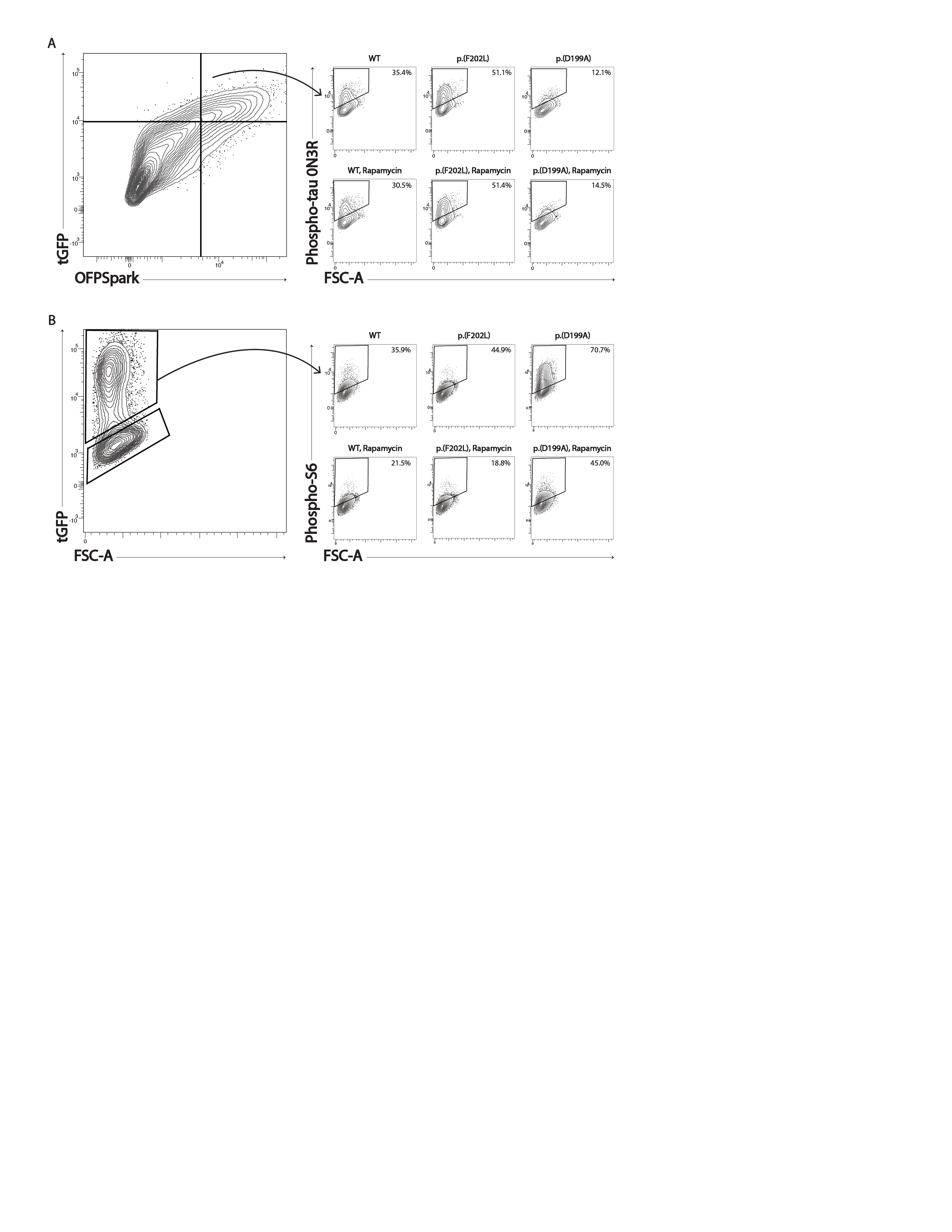

**Supplementary Figure 3 Contour plots for rapamycin experiments. (A)** Contour plots showing phospho-tau in control and rapamycin-treated tGFP^+^OFPSpark^+^ cells expressing MARK4^wt^, the p.F202L variant, and the p.D199A variant. **(B)** Contour plots showing phospho-S6 in control and rapamycin-treated tGFP^+^ cells expressing MARK4^wt^, the p.F202L variant, and the p.D199A variant. FSC: forward scatter; tGFP: turbo green fluorescent protein.
